# Differences of blood pressure values between Korotkoff and oscillometric device

**DOI:** 10.1101/2025.04.07.25325426

**Authors:** Chen Xing, Xu Jinsong, Su Hai

## Abstract

**Subject:** Using intro-aortic BP as reference,to compare the accuracy of a Korotkoff sound electronic device (Hanvon FY730) and an oscillometric device (Omron HBP-1320T3) in BP measurement,

**Methods:** BP was measured in catheterization coronary angiography. A crossover design was adopted for the using sequence of the two kinds of devices. The average of two BP readings were recorded as the final SBP, DBP and mean arterial pressure (MAP). Non-invasive BP measurement was initiated when the intro-aortic BP curve was stabilized, and the intro-aortic BP curve was simultaneously recorded for 15–20 seconds. BP differences (ΔBP) between non-invasive and its relative intro-aortic BP values were calculated as Oscillometric BP – intro-aortic BP difference (Osc-Ao) or Korotkoff sound method – intro-aortic BP difference (Kor-Ao).

**Results:** Using intro-aortic BP as the reference, the Korotkoff sound device provided closer readings compared to the oscillometric device.The Kor-Ao SBP was 19.5 ± 17.6 mmHg, but Osc-Ao SBP was 24.8 ± 20.2 mmHg (P < 0.001). Similarly, the MAP difference was smaller for the Korotkoff device (10.5 ± 10.9 mmHg vs. 13.7 ± 11.3 mmHg, P = 0.007). Although the DBP difference was also smaller for the Korotkoff device (6.1 ± 12.3 mmHg vs. 8.1 ± 11.5 mmHg), but no statistical significance (P = 0.147).

**Conclusion:** Using intro-aortic BP reading as reference, the Korotkoff sound device provides more accurate brachial SBP, DBP and MAP values against the oscillometric device.

---

The Korotkoff sound method was invented by Dr Korotkoff in 1905. The principle of mercury column-auscultation blood pressure(BP)measurement is based on this method. During mercury column-auscultation BP measurement, when the cuff pressure decreases and blood begins to flow through the brachial artery, the first Korotkoff sound means systolic BP (SBP); as the cuff pressure further decreases, the IV sound or V sound mean diastolic pressure (DBP). At present, column-auscultation BP measurement remains the gold standard for the verification of non-invasive BP device [1]. To avoid mercury pollution, electronic manometers become to replace mercury columns, but, the Korotkoff method can be fully utilized. In fact, Korotkoff sound electronic device has already been marketed domestically in China [2].

Even the oscillometric electronic devices are currently widely used in clinical practice, the controversy on this BP device is still continuous. The primarily of oscillometric method relies on the correlation between pulse waves (OPP) in the cuff and cuff pressure. An envelope curve is constructed based on this relationship, where the peak of the envelope curve coincides with the mean arterial pressure (MAP), and the corresponding cuff pressure at this time may be the same as the MAP value. Systolic and diastolic pressures(SBP and DBP)are then calculated using a relatively fixed formula [3], though different devices employ distinct calculation algorithms.

The objective of this study is to compare the accuracy of a Korotkoff sound electronic device (Hanvon FY730) and an oscillometric device (Omron HBP-1320T3) in BP measurement, using BP in aortic root (intro-aortic BP) as reference.

## 1 Subjects and Methods

The study protocol and informed consent procedures were approved by the Ethics Committee of the Second Affiliated Hospital of Nanchang University. All participants provided written informed consent.

### 1.1 Subjects

A total of 38 inpatients with the symptom of chest pain, who need cardiac catheterization coronary angiography were enrolled between February and March 2025. Exclusion criteria included: acute myocardial infarction; aortic coarctation; congenital heart disease; acute heart failure; hemiplegia; pulseless disease; prior transradial procedure and arrhythmia.

### 1.2 BP Measurement

BP measurements were conducted in the cardiac catheterization laboratory for the patients for coronary angiography.

#### 1.2.1 Non-Invasive BP measurement

Non-invasive BP was taken on the left upper arm. A crossover design was adopted for the sequence of using the two kinds of devices. In Group A, the device sequence was: Korotkoff sound device → oscillometric device → Korotkoff sound device → oscillometric device. In Group B, the sequence was reversed: oscillometric device → Korotkoff sound device → oscillometric device → Korotkoff sound device. The average of two BP readings were recorded as the final SBP, DBP and mean arterial pressure (MAP), meanwhile, the pulse rate (representing heart rate) was recorded.

#### 1.2.2 BP in aortic root

Patients were in supine position on the operating table. A 5Fr or 6Fr arterial sheath was inserted into the right radial artery to locate the tip of a Judkins catheter in the aortic root.

#### 1.2.3 Comparison of non-invasive and intro-aortic BP

Non-invasive BP measurement was initiated when the intro-aortic BP curve was stabilized. At the same time, the intro-aortic BP curve was simultaneously recorded for 15–20 seconds. Each non-invasive BP reading had relatively intro-aortic BP reading as reference. In this study, the average of two non-invasive BP readings was used for analysis [4].

#### 1.2.4 Intro-aortic and non-invasive BP differences

BP differences (ΔBP) between non-invasive and its relative intro-aortic SBP, DBP and MAP values were calculated, separately :

Oscillometric BP – intro-aortic BP difference (Osc-Ao);

Korotkoff sound method – intro-aortic BP difference (Kor-Ao).

##### Statistical Analysis

Data were entered into Excel 2003 and analyzed using SPSS 10.0. Continuous variables are expressed as mean ± standard deviation. Statistical analyses included t-tests, paired-sample t-tests, analysis of variance (ANOVA), and omnibus tests. Linear regression analysis was performed to examine the correlation between invasive and non-invasive blood pressure (BP) measurements. Bland-Altman analysis was used to evaluate the agreement between non-invasive blood pressure measurements and aortic root BP [10].

Bland-Altman plots were generated using Origin software, the x-axis represents the mean of non-invasive and invasive BP values and the y-axis represents the difference between non-invasive and invasive BP values. Scatter plots were created for non-invasive SBP, DBP and MAP against the corresponding intro-aortic BP values.The 95% limits of agreement (LoA) were determined (95% LoA = mean difference ± 1.96 × standard deviation). A p-value < 0.05 was considered statistically significant.

## 2 Results

### 2.1 General Characteristics of Participants

The general characteristics of the 38 study patients are presented in Table 1.

**Table 1.** General Characteristics of the Study Participants.

| Items | n=38 |
| --- | --- |
| Age [$\bar{x} \pm S, y$] | 63.9±10.9 |
| male[n ( % ) ] | 26 ( 68.4 ) |
| Hypertension [n ( % ) ] | 23 ( 60.5 ) |
| hyperlipoidemia [n ( % ) ] | 13 ( 34.2 ) |
| diabetes mellitus; [n ( % ) ] | 10 ( 26.3 ) |
| Coronary artery disease [n ( % ) ] | 33 ( 86.8 ) |
| LDL [mmol/L] | 2.8±0.9 |
| HDL [mmol/L] | 1.3±0.3 |
| TG [*mmol/L*] | 1.6±0.8 |
| TC [mmol/L] | 4.9±1.1 |
| ALT [u/L] | 60.8±164.4 |
| AST [u/L] | 53.4±123.0 |
| Creatinine [μmol/L] | 92.0±42.8 |
| eGFR [ml/min/1.73m<sup>2</sup>] | 81.5±25.7 |
| Glycosylated hemoglobin ( % ) | 6.0±0.6 |

### 2.2 Correlation analysis with intro-aortic BP

Regardless off the A or B groups, the intro-aortic BP values relative to the Korotkoff sound device or the oscillometric device were similar. In this study the correlation coefficients between intro-aortic SBP DBP and MAP relative to Korotkoff device or to oscillometric device were 0.984, 0.894, and 0.930, respectively (all p < 0.001) (Table 2).

**Table 2.** Intro-aortic BP relative to Korotkoff or oscillometric method in the 38 patients (mmHg)

|  | SBP | DBP | MAP |
| --- | --- | --- | --- |
| Intro-aortic BP relative to Korotkoff method | 122.1±19.7 | 73.3±13.5 | 89.6±12.7 |
| intro-aortic BP relative to oscillometric method | 122.3±19.5 | 73.5±14.1 | 89.8±13.1 |
| Correlation coefficient | 0.984 | 0.894 | 0.930 |
| P value | <0.001 | <0.001 | <0.001 |

### 2.3 Comparison of non-invasive and aortic BP differences

Using intro-aortic BP as reference, the Korotkoff sound device provided closer readings compared to the oscillometric device.The SBP difference was 19.5 ± 17.6 mmHg for the Korotkoff device. But those were 24.8 ± 20.2 mm Hg for the oscillometric device (P < 0.001). Similarly, the MAP difference was smaller for the Korotkoff device (10.5 ± 10.9 mmHg vs. 13.7 ± 11.3 mmHg, P = 0.007). Although the DBP difference was also smaller for the Korotkoff device (6.1 ± 12.3 mmHg vs. 8.1 ± 11.5 mmHg), but no statistical significance (P = 0.147) (Tables 3-1, 3-2 and 3-3).

**Table 3.**
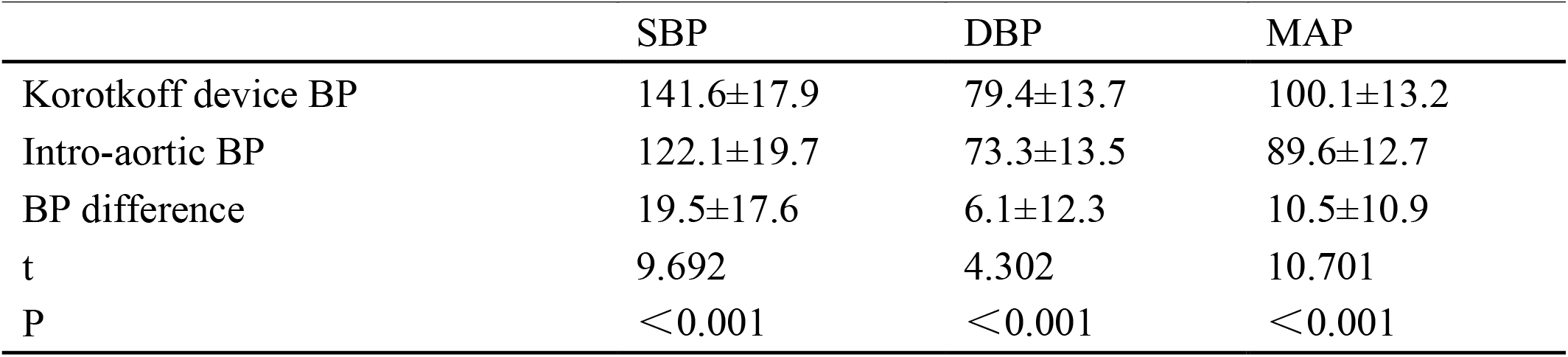
Korotkoff BP and the corresponding intro- aortic BP in 38 patients(mmHg)

**Table 4.** Oscillometric BP and the corresponding intro-aortic BP in 38 patients (mmHg)

|  | SBP | DBP | MAP |
| --- | --- | --- | --- |
| Oscillometric device BP | 147.1±19.6 | 81.6±11.5 | 103.5±12.9 |
| Intro-aortic BP | 122.3±19.5 | 73.5±14.1 | 89.8±13.1 |
| BP difference | 24.8±20.2 | 8.1±11.5 | 13.7±11.3 |
| t | 6.108 | 8.463 | 10.55 |
| P | <0.001 | <0.001 | <0.001 |

**Table 5.** Comparison of the difference between Korotkoff - intro-aortic or oscillometric and intro-aortic BP (mmHg)

|  | ΔSBP | ΔDBP | Δ MAP |
| --- | --- | --- | --- |
| Korotkoff - intro-aortic | 19.5±17.6 | 6.1±12.3 | 10.5±10.9 |
| Oscillometric - intro-aortic | 24.8±20.2 | 8.1±11.5 | 13.7±11.3 |
| BP difference | -5.3±12.0 | -2.0±12.1 | -3.1±9.8 |
| t | -3.886 | -1.464 | -2.775 |
| P | <0.001 | 0.147 | 0.007 |

## 4 Bland-Altman Analysis

Bland-Altman analysis demonstrated that the differences between brachial SBP and MAP from the Korotkoff sound device and intro-aortic relative BP values were smaller compared to those of the oscillometric device. This result supports the superior agreement of the Korotkoff sound device with invasive aortic measurements for SBP and MAP.

Additionally, the 1.96 SD ranges for SBP and MAP by the Korotkoff device were narrower: SBP: Korotkoff device : -14.8 to 53.9 mmHg; oscillometric device : -14.8 to 64.5 mmHg; DBP: Korotkoff device : -18.0 to 30.2 mmHg; oscillometric device : -14.5 to 30.7 mmHg; MAP: Korotkoff device : -10.7 to 31.8 mmHg; oscillometric device : -8.5 to 35.8 mmHg (Figure 1).

**Figure 1.**
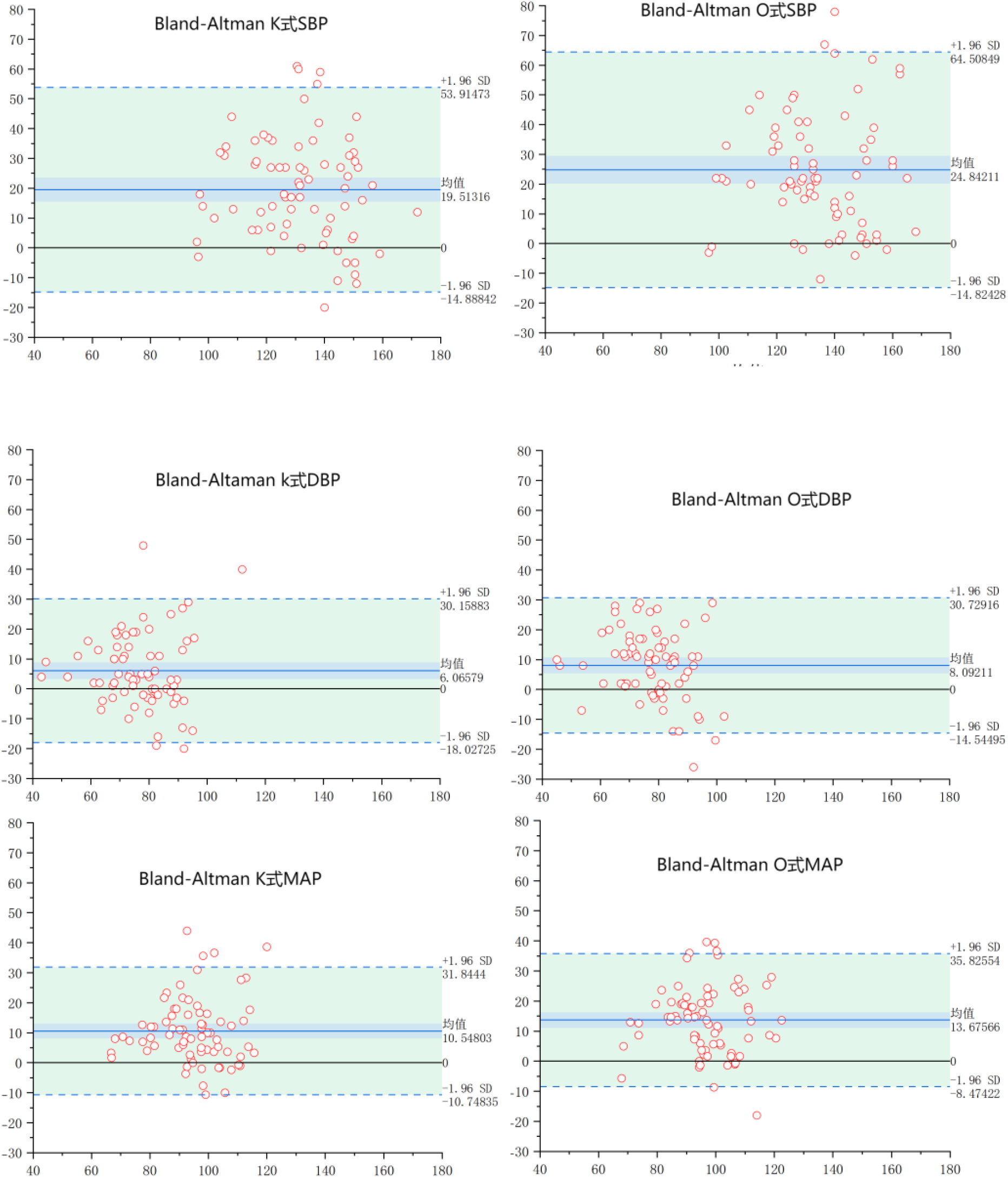
Bland-Altman analysis of SBP, DBP and MAP K: Korotkoff device ; O: Korotkoff device

## 3 Discussion

Among the 38 patients underwent coronary angiography, BP of left upper arm was measured twice using a Korotkoff sound device (Hanvon FY730) and an oscillometric device (OmronHBP-1320T3). Each non-invasive BP reading was paired with a relative intro-aortic BP reading, by this way to ensure accurate <u>comparison</u>. Consequently, 152 paried non-invasiveand and intro-aortic BP values were obtained from the 38 patients.

The traditional stethoscope is replaced by an acoustic sensor, meanwhile, this device incorporates innovations in design, hardware, data processing, and algorithmic software. At firs, the cuff pressure is initially increased to 30 mm Hg above SBP. As the cuff pressure gradually decreases, blood flow resumes in the brachial artery, generating the first Korotkoff sound (detected electronically), the relative cuff pressure recorded as SBP. Further deflation leads to the disappearance of vascular sounds (typically Phase V Korotkoff sound), with the pressure at this point defined as DBP.

The study revealed that the differences between Korotkoff-Ao SBP, DBP and MAP measured were significantly smaller than those obtained with the oscillometric device (Oscillometric-Ao). In other words, the results indicate that using intro-aortic BP as the reference, the Korotkoff sound device provides more accurate SBP, DBP and MAP as compared to the oscillometric device. In our study, SBP values from the Korotkoff device were closer to intro-aortic values by 5.3 mmHg; for DBP by 2.0 mmHg; amd for MAP by 3.2 mmHg.

Recently, several kinds of automated auscultatory upper-arm cuff devices were developed in other counties, such as the InBody BPBIO480KV [6], KOROT V2 Doctor (InBody BPBIO280KV), and KOROT P3. The principle of these devices was similar to the device used in this study. These devices have passed the validation trials compliance with the AAMI/ESH/ISO Universal Standard and were used in clinical [7-9].

Ar present, the mercury column-auscultation method remains as the gold standard for validation of non-invasive electronic devices. In this study, using intro-aortic BP as the reference, the Korotkoff sound electronic device provides more accurate SBP, DBP and MAP than the oscillometric device. For the reason of the difference, we suggested that the key critique of oscillometric devices is their reliance on “one-size-fits-all” formulas to calculate SBP and DBP among different individuals with varying ages, genders and body weights. This simple approach may lead to significant individual BP discrepancies and partial inaccuracies. In contrast, the Korotkoff sound method enables personalized BP readings by directly detecting vascular sounds, thereby capturing individualized hemodynamic profiles.

### Clinical Significance

This is the first study to compare arm BP readings between Korotkoff sound electronic device and an oscillometric device using the intro-aortic BP as the reference. Among the 38 patients underwent coronary angiography and when the intro-aortic BP levels relative to the Korotkoff device or to the oscillometric method were similar, the Korotkoff electronic BP device provide closer SBP, DBP and MAP. These findings confirm the superior accuracy of the Korotkoff device against the oscillometric device.

### Limitation

The intro-aortic BP levels in the 38 study patients were relatively low, because a well-controlled BP was need for coronary angiography. Whether the results of this study represent systematic consistency in patients with higher BP remains uncertain, further data accumulation is needed. Additionally, larger-scale studies is needed in the future.

## Conclusion

Using intro-aortic BP reading as reference, the Korotkoff sound BP device provides more accurate SBP, DBP and MAP values against the oscillometric device.

## Data Availability

the availability of all data referred to in the manuscript

